# The impact of ancestry on performance of type 1 diabetes genetic risk scores: high discrimination performance is maintained in African ancestry populations, but population specific thresholds may improve risk prediction

**DOI:** 10.1101/2025.07.17.25330632

**Authors:** Steven Squires, Jean Claude Katte, Dana Dabelea, Catherine Pihoker, Jasmin Divers, Eugene Sobngwi, Moffat J. Nyirenda, Raymond J. Kreienkamp, Angela D Liese, Amy S Shah, Lawrence Dolan, Kristi Reynolds, Maria J. Redondo, William Hagopian, Segun Fatumo, Mesmin Y. Dehayem, Andrew Hattersley, Michael N. Weedon, Angus Jones, Richard A. Oram

## Abstract

**OBJECTIVE:** Genetic risk scores (GRSs) for type 1 diabetes (T1D) may assist T1D classification and prediction but are often developed from European populations. To improve health outcomes, it is important to understand the performance and utility of GRSs in diverse ancestry populations.

**RESEARCH DESIGN AND METHODS:** We assessed performance of three previously published T1D GRSs in differentiating people with and without Type 1 diabetes in African (with/without T1D=194/235), European (n=1109/125), and Hispanic (266/170) ancestry populations in the USA, and from Cameroon and Uganda (n=144/5001). The assessed GRSs were developed from European ancestry populations (GRS1, GRS2) and from an African ancestry population (AAGRS).

**RESULTS:** The discriminative power, as measured by the area under the receiver operating characteristic curve (AUC), for GRS2 and AAGRS were equivalent on the African ancestry populations, and both outperformed the GRS1: the AUCs produced by the GRS2, AAGRS and GRS1 on Uganda/Cameroon data were 0.882 (0.845-0.914), 0.874 (0.838-0.907) and 0.816 (0.772-0.857) respectively. GRS2 outperformed the AAGRS and GRS1 on Hispanic and European populations. The GRS2 distributions varied by population, with lower average scores for African populations. If the same GRS2 risk thresholds of 11.5 were set for European and African populations, the sensitivities were 0.91 and 0.53, respectively.

**CONCLUSIONS:** The GRS2 produced similar or improved discriminative power across the populations but the AAGRS matched performance on African ancestry participants with fewer single nucleotide polymorphisms. Varying GRS2 risk thresholds may be required for different populations due to the divergent distributions.

## INTRODUCTION

Genetic risk scores (GRSs) summarise an individual’s known genetic variation about the risk of developing a disease into a single number. There is considerable interest in the use of GRSs for stratification of populations into risk classes to enable improved clinical outcomes (1), for example, through targeted monitoring of higher risk groups. GRSs can also be combined with other measures of risk to improve prediction or classification models (2), and can assist with study of diabetes aetiology (3). GRSs perform particularly well in type 1 diabetes (T1D) due to the high heritability of T1D, explained by a small number of variants in the human leukocyte antigen (HLA) region (4,5).

GRSs are derived from genome wide association studies (GWASs), which find associations between individual single nucleotide polymorphisms (SNPs) and risk of a disease. These produce statistical significance of the SNPs alongside weights for use in a linear model. GWASs are predominantly performed in Europeans (6) with fewer studies in people of other ancestries (7). To reach SNP-wise statistical significance requires considerable quantities of genetic data so GRSs are either not produced, or may capture less genetic risk, in groups underrepresented in GWASs. An alternative to generation of GRSs for different populations is to utilise previously developed GRSs but the transferability of GRSs is often poor (8) or unknown. As GRSs become more useful for medicine (9) and research (10) we may see further increases in health disparities if the quality and validation of GRSs for underrepresented groups is not improved.

T1D is less well studied and therefore the aetiology less well understood in people of African ancestry than in European populations (11). Recent analyses have shown genetic overlap in T1D between European and African ancestry populations (12–15). To date several GRSs have shown good discriminative ability in African-Americans (12,13), but there are no tests of these scores in populations in Africa. In this study we investigated how two European derived GRSs (GRS1 (16) and GRS2 (17)), and one African GRS (AAGRS (13)) performed on a dataset from the USA of people living with T1D and people without T1D which includes people of European, Hispanic, and African ancestry alongside a dataset with T1D and non-T1D from Cameroon and Uganda in sub-Saharan Africa.

## RESEARCH DESIGN AND METHODS

We applied two T1D GRSs discovered on European populations (GRS1 and GRS2) and one T1D GRS discovered on an African ancestry population (AAGRS) on five datasets consisting of people with T1D and those without T1D. Three of these datasets contain participants with African ancestry with one group from the USA, one from Cameroon and one from Uganda. The other two groups are Europeans and Hispanics from the USA. We then investigated the performance of the three GRSs on these five datasets with the primary focus on the capacity of the GRSs to discriminate between T1D and non-T1D participants; we also examined effect of ancestry on risk thresholds; and how different ancestry populations in the same study would affect the apparent performance of the GRS. Furthermore, we investigated relationships between the GRSs, whether performance improved by combining them, and the importance of individual SNPs to the risk estimates.

### Research Cohorts

#### USA

Data from the USA came from The SEARCH for Diabetes in Youth study (18), an epidemiology study which includes people of African, European and Hispanic ancestries, labelled US-Africans, US-Europeans and US-Hispanics respectively, with predominantly T1D or type 2 diabetes (T2D) diagnosed before age 20. In this study we defined T1D as a clinical diagnosis alongside at least one autoantibody (of GAD, IA2 and Znt8) and non-T1D as those with a clinical diagnosis of T2D. We defined ancestry by clustering in the first two principal components of the genetic data. For US-Africans there were 194 participants with T1D and 235 without; for US-European there were 1109 participants with T1D and 125; and for US-Hispanics 266 with T1D and 170 without. These numbers for each ancestry and T1D status are additionally recorded in Table S1 of the supplementary material.

### Uganda and Cameroon

The sub-Saharan ancestry T1D cohorts come from the Young Onset Diabetes in sub-Saharan Africa (YODA) study (3) (Clinical ID: NCT05013346) which includes participants from Cameroon and Uganda diagnosed with T1D before age 30. The T1D cases are defined as a clinical diagnosis of T1D alongside at least one positive autoantibody (of GAD, IA2 and ZNt8). Non-T1D data is from population data for Uganda (19) and Cameroon (20). We show results of these two datasets combined, with separate results in the supplementary material. The number of Uganda and Cameroon participants with T1D was 144 and those without was 5001. Details of these datasets are also in supplementary Table S1.

### 1000G

To improve confidence in our conclusions we also used additional data from those without T1D from the 1000G project (21) which was an international study aiming to catalogue diverse genomes. We used African and European non-T1D samples from the 1000G to compare with the African and European ancestry cohorts; for comparison with the Hispanic population, we used the Americas superpopulations. Details of this data are in supplementary Table S1 (labelled as Non-T1D 1000G).

### Genotyping, quality control, and imputation

The SEARCH genotyped data was produced on two chips: the Multi-Ethnic Global Array (MEGA; Illumina) and the Affymetric 500k imputation scaffold chip. We performed quality control, detailed in the supplementary material, on the data from the separate chips before merging them, removing any non-common SNPs and imputing using the TOPMed imputation panel (22) and the Minimac server (23,24).

The YODA T1D data from Cameroon and Uganda was genotyped on a Global Screening Array (GSA; Illumina) chip. The data was quality controlled together and imputed using TOPMed. The Cameroon controls and Uganda population data were quality controlled separately and imputed to TOPMed.

For the GRS2, some of the required SNPs could not be accurately imputed, primarily due to very low allele frequencies. The effect of these poorly imputed SNPs is discussed in the supplementary material. The overall effect is likely to be slightly poorer discriminative power but with no impact on the conclusions drawn.

### GRSs

Two GRSs developed on European populations were calculated. The first (16) (denoted here as GRS1) includes 30 SNPs, with 28 contributing a linear weighted sum of the effect alleles with the weights produced from a GWAS. The final two SNPs are used in combination to tag the DR3/DR4-DQ8 haplotypes. The second European GRS (denoted as GRS2) (17) contains 67 SNPs of which 14 tag DR-DQ haplotypes, with specific DR-DQ interaction terms for 18 haplotype combinations. The other 53 SNPs are included in the linear weighted sum with 21 from other SNPs in the HLA and 32 from the non-HLA. This second European GRS has shown higher performance than the GRS1 and is believed to more comprehensively capture HLA and non-HLA risk. We additionally calculated a version of the GRS2 with a reduced number of SNPs (denoted as GRS47); with results available in the supplementary material.

We also calculated a GRS (AAGRS) developed in people of African ancestry (13), which contains 7 SNPs with 5 in the HLA and 2 in the non-HLA. The AAGRS was shown to perform statistically significantly better than GRS1 in African ancestry populations (13).

### Statistical Methods

We analysed the discriminative performance of the three GRSs at separation of the T1D from non-T1D participants in the African ancestry cohorts (SEARCH, Uganda and Cameroon) as well as the European and Hispanic ancestry participants from SEARCH by considering the receiver operating characteristic (ROC) curves and the area under the ROC curves (AUC). The 1000G data was utilised to assess similarities in distributions between the non-T1D data.

We also explored how the sensitivity (true positive rate) and specificity (true negative rate) would alter by ancestry with choices of different thresholds to separate the populations into high and low risk groups. In addition, we explored how conclusions about overall GRS performance and the general separability of the data would change if we had different populations within the same dataset.

The similarity of the GRSs on the datasets was investigated using correlation coefficients. We also examined if overlapping genetic information from the AAGRS and GRS2 could be combined for better performance. For the AAGRS we investigated how pairwise combinations altered performance. For the AAGRS and GRS2 we explored how the differences in average weighted scores of the SNPs contributed to the final differences between T1D and non-T1D, as well as adding all possible combinations of the 7-SNPs from AAGRS to GRS2. We also assessed which SNPs were important to the AAGRS and GRS2 discrimination performance.

## RESULTS

### Discriminative power of GRS1, GRS2 and AAGRS on the different populations

We demonstrate the varying performance of the three GRSs on the datasets in Figure 1. GRS2 and AAGRS showed similar performance in Africans and those of African ancestry; the AUCs for GRS2 and AAGRS on Uganda/Cameroon were 0.882 (95% CI 0.845-0.914) and 0.874 (95% CI 0.838-0.907), respectively; while the AUCs for GRS2 and AAGRS on the US-Africans were 0.839 (95% CI 0.799-0.877) and 0.838 (95% CI 0.799-0.874) respectively. The GRS1 had a statistically significantly reduced performance compared to GRS2 or AAGRS on the Uganda/Cameroon and US-Africans populations with AUCs of 0.816 (95% CI 0.772-0.857) and 0.796 (95% CI 0.751-0.837), respectively. In the US-European data, the GRS2 produced an AUC of 0.878 (95% CI 0.841-0.912), which significantly outperformed GRS1 and AAGRS with AUCs of 0.840 (95% CI 0.799-0.876) and 0.810 (95% CI 0.764-0.851), respectively. Similarly, for US-Hispanic data the GRS2 produced an AUC of 0.868 (95% CI 0.832-0.901), which was significantly higher than GRS1 or AAGRS, which had AUCs of 0.807 (95% CI 0.767-0.856) and 0.789 (0.742-0.834), respectively. In Table S2 of the supplementary material, we show the AUCs with uncertainty estimates for these results and for the separate Uganda and Cameroon data. In supplementary Figure S2, we show separate ROC curves for the Uganda and Cameroon populations. In supplementary Figure S3 we show ROC curves and AUCs for the GRS47 as well as plots of the GRS47 against GRS2; findings for GRS47 were similar to GRS2. We also used the 1000G data as non-T1D data showing similar discrimination performance by AUC (supplementary Table S3).

**Figure 1.**
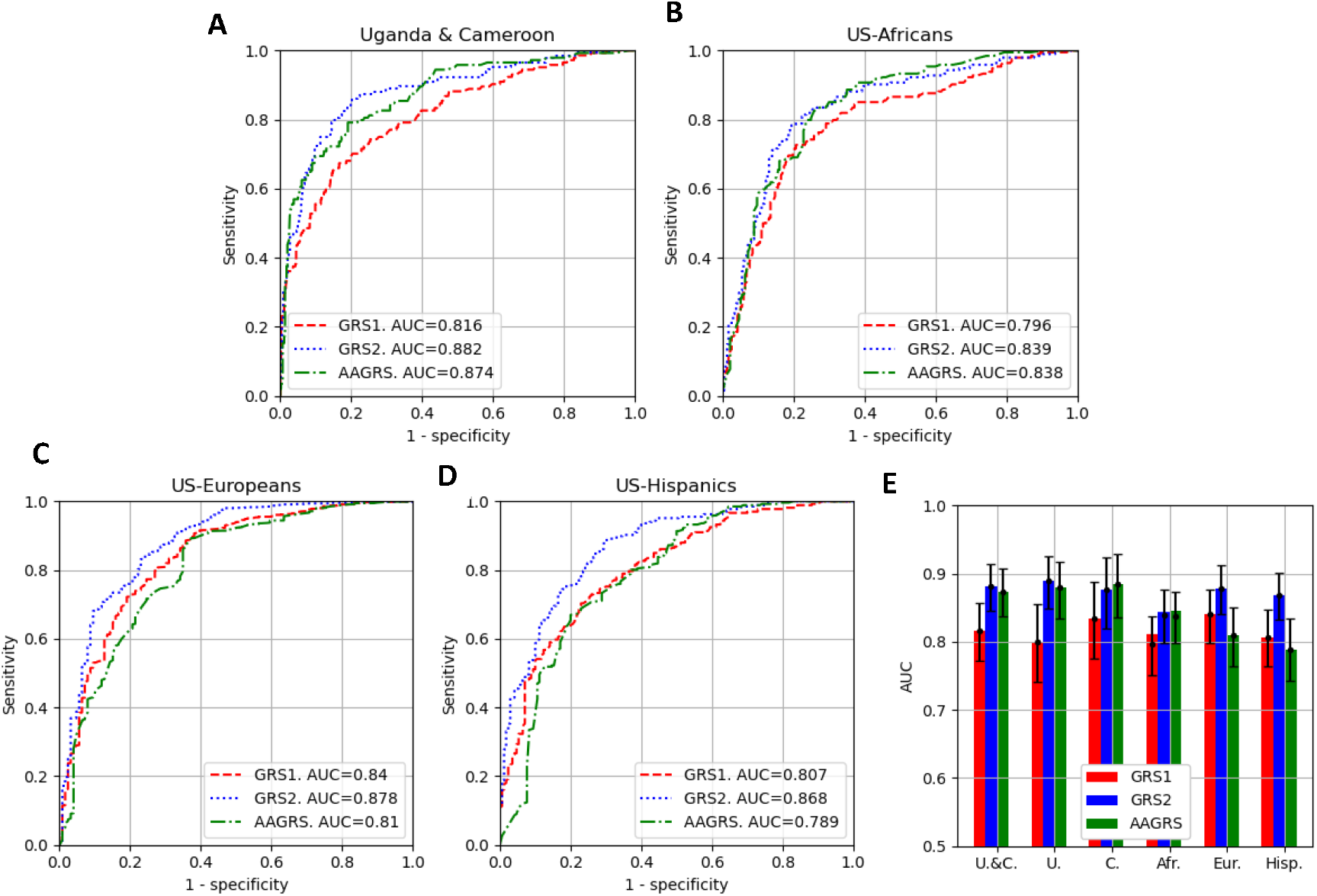
The discriminative performance of the three GRSs (GRS1, GRS2, AAGRS) at separation of the T1D from non-T1D for the datasets. A, B, C, D: ROC curves for the three GRSs and the populations. Associated AUCs are shown in the legend. E: The AUCs with 95% uncertainties for the three GRSs and the populations Uganda and Cameroon combined (U.&C.), Uganda (U.), Cameroon (C.), US-Africans (Afr.), US-Europeans (Eur.) and US-Hispanics (Hisp.).

### Effect of ethnicity on GRS1, GRS2 and AAGRS distributions

The GRS distributions for the three GRSs on the datasets are shown in Figure 2. The African ancestry datasets had a lower distribution of scores for both non-T1D and T1D participants than the equivalents for the Europeans and Hispanics, which holds for all three GRSs. The Uganda and Cameroon data are shown separately in supplementary Figure S4. In supplementary Figure S5 we show distributions of two risk scores calculated by splitting the GRS2 into HLA (35 SNPs) and non-HLA (32 SNPs); most discrimination power came from the HLA SNPs but the change in GRS distribution was driven by both HLA and non-HLA SNPs. We also show the distributions of the GRSs on the 1000G data in Figure S6 of the supplementary material for superpopulations associated with African, Hispanic and European populations.

**Figure 2.**
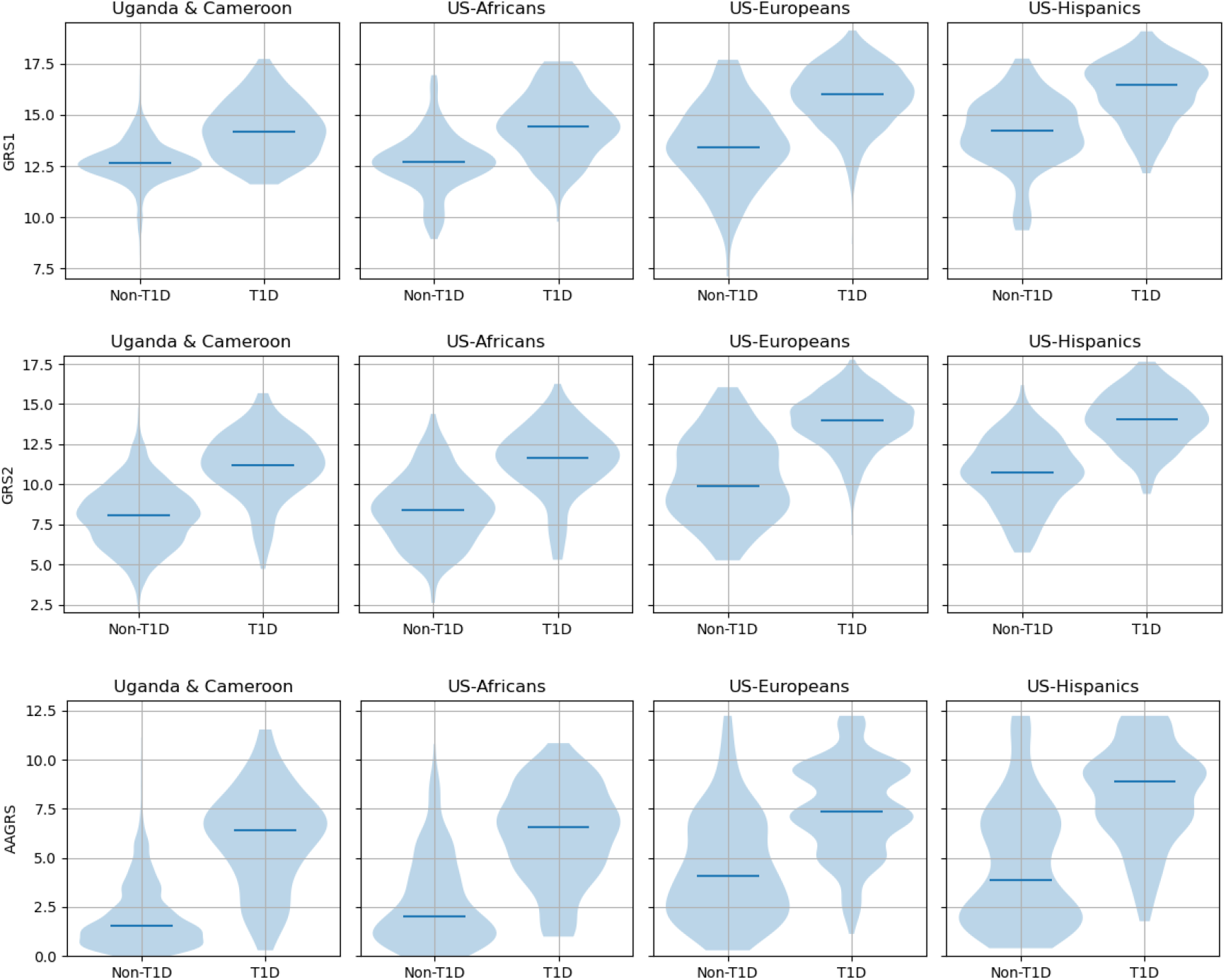
GRS distributions for the three GRSs and the T1D and non-T1D populations.

### Sensitivity and specificity with different risk thresholds

In Figure 3 (A, B) we show how, for GRS2, the sensitivity and specificity index vary with the choice of threshold defining those at lower and higher risk. The top left (A) and right (B) plots show the sensitivity and specificity respectively against GRS threshold for the datasets. The three African ancestry populations show similar patterns, which differ from the European or Hispanic, with generally lower sensitivity and higher specificity for the same threshold.

**Figure 3.**
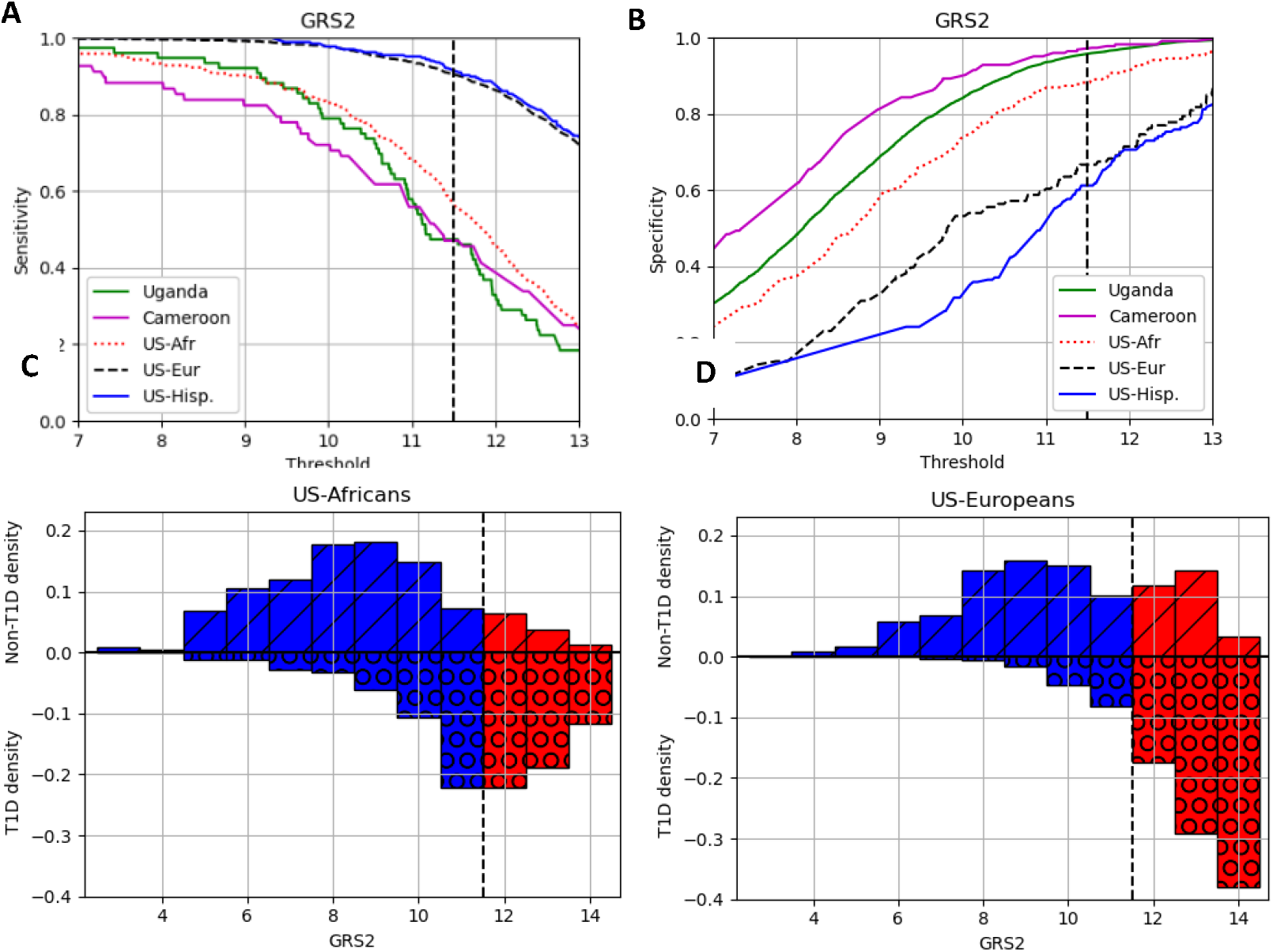
The importance of risk thresholds for the GRS2 in the different populations. A, B) The sensitivity and specificity against risk threshold, respectively. The sensitivity/specificity is shown per population for each of the risk threshold values. The dashed vertical line shows the 11.5 threshold for generating the other plots. C and D) the densities of the non-T1D and T1D data at different GRS values. Samples to the left of the vertical dashed line would be defined as low risk (in blue columns) and to the right of the line as high risk (red columns). The number of non-T1D and T1D samples are shown as bars above (and marked with diagonal dashes) and below (marked with circles) the horizontal line, respectively.

If the same threshold was chosen, there would be differences in both sensitivity and specificity for the populations. We demonstrate this effect of choosing one threshold (at 11.5) for the SEARCH African (bottom left plot, C) and European ancestry (bottom right plot, D). For the US-Africans there are small numbers of non-T1D being defined as high risk while for the US-Europeans there are more, i.e., the specificity of the US-Africans is higher than the US-Europeans. Conversely, for the US-Africans many of the US-Africans T1D samples are (falsely) defined as low risk while for the US-Europeans only few are. Equivalent plots for the other datasets are shown in the supplementary material (Figure S7). We also show confusion matrices in the supplementary material (Figure S8) demonstrating this same effect.

### The importance of correctly matching populations for cases and controls

In Figure 4A we demonstrate the effect of assessing the quality of the GRS2 with different populations. We calculated the AUC produced if each of the five populations were assessed compared to each other for cases and controls. We then removed the expected AUC if the population cases/controls were correctly compared to itself; the expected AUC is the AUC between the cases and controls for the same population.

**Figure 4.**
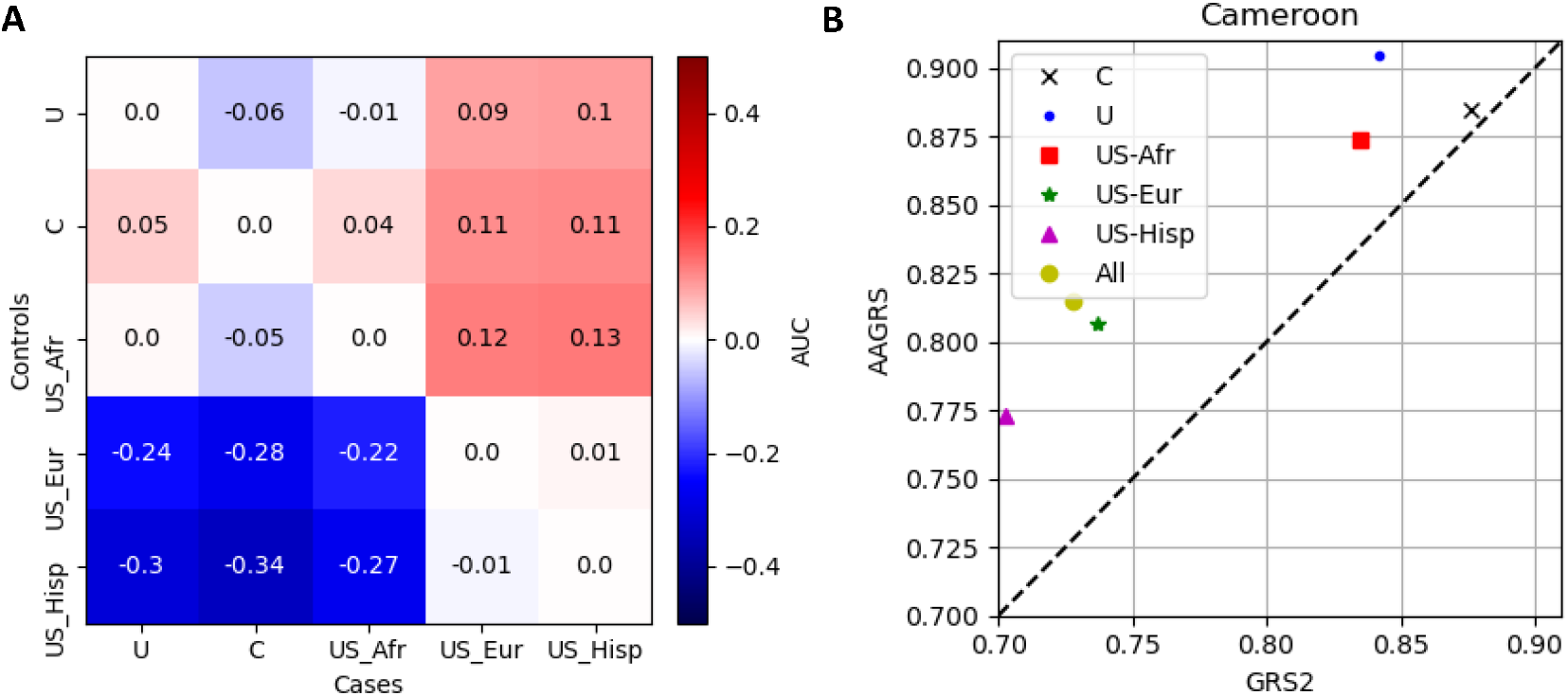
AUCs for the GRS2 if comparing different populations. The heatmap (A, left plot) shows the differences in AUC from the expected (defined as the AUC between data from the same population) if comparing different populations. Negative values mean the AUC would appear lower and positive values the AUC would appear higher. The right plot (B) shows the AUCs for the AAGRS and GRS2 if the non-T1D controls combine populations (with under-sampling so the non-T1D population importance is equalised) for the Cameroon T1D. The black cross is where the non-T1D population is just Cameroon, the yellow circle is for all five populations included in the non-T1D populations, and the others are combinations of Cameroon with one other population.

If the value is below 0 then the AUC between those pair of populations is lower than would be expected. For examples the largest gap is between the T1D Cameroon (C) and the US-Hispanic non-T1D which has a gap of -0.34. Conversely, the AUC would be overstated if the value is positive, so for the comparison of T1D US-Europeans with non-T1D US-Africans the apparent AUC of the comparison would be 0.13 higher than it should be. For all these plots, the pattern is the same with the comparisons between the US-Europeans or US-Hispanics with any of the three African populations yielding apparently higher discriminative power. Conversely the comparison between any of the three African populations with the US-Europeans or US-Hispanics would yield apparently worse performance.

Another scenario is the comparison of a population with a set of mixed populations, in Figure 4B we show how this would alter the apparent quality between the GRS2 and AAGRS. We used the Cameroon T1D samples and calculated the AUC for the GRS2 and AAGRS if the non-T1D data was a combination of the Cameroon data with the other four datasets, both individually (the Cameroon non-T1D data and one other dataset) and for all datasets (the Cameroon data with all other four datasets). We under-sampled from the larger non-T1D datasets so we had the same weighting to the non-T1D data. The labels in the legend show which other non-T1D results are included with the Cameroon data. When the Cameroon non-T1D data is used on its own (black cross) the AAGRS and GRS2 show similar discrimination performance. When including the other datasets there is a change in apparent quality of the AAGRS and GRS2, with the GRS2 appearing to perform worse. Complementary plots for the other datasets are shown in supplementary Figure S9.

### Direct comparisons of GRSs and combining GRSs

The direct relationships between the GRSs are shown in Supplementary Material with comparisons of the three GRSs for the five populations shown in Figure S10. The Pearson correlation coefficients for the combined T1D and non-T1D scores, are shown in Table S4; the correlations between GRSs tended to be higher in the SEARCH than the YODA data.

We also considered the effect on the AUCs by combining together the final GRS2 and AAGRS. In supplementary Figure S11 we show all possible combinations of the weighted SNPs from the AAGRS with the GRS2 final score. The histogram shows the distribution of the scores, with the dashed, dotted and solid lines showing AUCs for the GRS2, AAGRS and final summed scores respectively. In the bottom right plot we show the individual AUCs, the sums (*Summed)* and when we weight the two scores to optimise performance (*OptSummed*). For the African ancestry populations the AUC can be higher by combining the GRSs, or adding some of the AAGRS SNPs to the GRS2, but the changes are not statistically significant. For the Hispanic and European populations adding SNPs or scores from the AAGRS to the GRS2 tends to reduce the performance of the GRS2, although not statistically significantly.

### SNP importance for the AAGRS and GRS2

For GRS2, we show the relative importance of the SNPs for the separation of classes by plotting the allele frequencies of the T1D against non-T1D for the datasets (supplementary Figure S12). The risk increasing and protective alleles are red crosses and blue dots, respectively, and the marker size is proportional to the effect magnitude. We also show, in supplementary Figure S13, similar plots for the three African ancestry populations compared to one another to demonstrate the similarities in allele frequencies. In Supplementary Table S6, we show those SNPs which have either an average (across the five populations) difference of over 0.1 or individual population differences over 0.1, of which there are 11.

For the AAGRS we consider the AUCs for the populations of individual, pairwise and all possible combinations of SNPs. The inclusion and removal of pairs of SNPs is shown in supplementary Figures S14. With just two of the 7-SNPs (rs2187668 and rs9273363) much of the discriminative power is still available.

We also consider the effect of individual SNPs on the GRS2 performance by calculating the differences between the average of the weighted SNPs for the T1D and the non-T1D samples for each population. In general, the larger these differences, are the more important the SNPs are to the capacity of the GRS to separate the T1D from non-T1D (accepting effects of linkage disequilibrium which may make some of the SNPs partially redundant). In supplementary Table S5 we show the 11 most important SNPs from the GRS2 for separation of T1D from non-T1D.

### Correlations between AAGRS and GRS2 SNPs

The GRS2 and AAGRS show a similar level of performance on the African ancestry datasets with different combinations of SNPs. In supplementary Figure S15 we show the correlations between the AAGRS SNPs and the previously mentioned 11 most important SNPs from the GRS2. There is substantial correlation between some of these SNPs.

## CONCLUSIONS

On the three African ancestry populations the discriminative power between people with T1D and those without T1D of the GRS2 and AAGRS was higher than the GRS1; there was no statistically significant difference in the AUCs between GRS2 and AAGRS. This suggests that GRS2 and AAGRS are capturing similar risk information for the people of African ancestry which was not captured by GRS1. The GRS2 outperforms both the AAGRS and GRS1 on the Hispanic and European populations.

The GRS2 either outperformed, or matched, the discriminative performance (defined by the AUC) of the GRS1 and AAGRS. Therefore, from this data, it could be used as an across-ancestry T1DGRS without loss of performance. A caveat is that GRS2 requires 67 SNP to be available rather than the 7 for the AAGRS and it is also a more complicated GRS with interaction terms between SNPs making it more difficult to accurately generate.

The distributions of the GRSs varied considerably between the datasets; the three African populations generally have lower levels of all three GRSs compared to the European and Hispanic populations, in both participants with and without T1D. If the GRS2 was used as an across-ancestry GRS, then there are several consequences for its use. One is that the choice of a risk threshold would need to be considered carefully for different populations. If the same threshold were used for different populations the sensitivity and specificity would be substantially different; the African ancestry populations had higher specificity and lower sensitivity than the European or Hispanic populations at the same risk threshold. A second issue is that the judgement about the quality of the model, judged by AUC, would need to consider ancestry of T1D cases and non-T1D controls to avoid drawing false conclusions about the quality of the discrimination. Imbalance in ethnicity of cases and controls could lead to either under-or over-estimating the performance of the GRS.

The correlation between the final scores of the three GRSs (on the same population) varies considerably, given the AAGRS and GRS2 produce similar discriminative performance on the African populations the correlations between these scores are modest. These differences in scores do not, however, result in any statistically significant improvement in AUC if the final scores are summed. For the three African populations the sum of the scores increases the AUC but not statistically significantly. The addition of the AAGRS scores to the GRS2 for the Europeans and Hispanics does not improve performance. Similarly, adding any combinations of the AAGRS weighted SNPs does not significantly improve the performance of the GRS2. Both these conclusions seem consistent – we would expect some modest improvement by adding the AAGRS as it would additionally emphasise African ancestry SNPs. However, the AAGRS and GRS2 both produce high AUCs and it is usually harder to improve an already effective classifier. The additional noise of extra SNPs to non-African ancestry data seems likely to slightly reduce the overall quality.

Within the GRS2 there are a modest number of SNPs that are particularly important when considering the differences in average weighted values. Across the five populations there is reasonable agreement about the importance of many of the SNPs; while there are some SNPs that are more important for different populations. These European derived significant SNPs appear to capture substantial risk information on the African ancestry populations. For the AAGRS only a few SNPs (especially two HLA SNPs rs2187668 and rs9273363) produce most of the discriminative power, although all seven have some power on their own.

The eleven most important GRS2 SNPs (defined by differences in mean scores between T1D and non-T1D) have substantial correlations with the AAGRS SNPs. There are complexities here with some differences between the populations but the overlap between the important GRS2 SNPs and SNPs from the AAGRS on the African population was substantial.

Overall, a European ancestry GRS (GRS2) performed well on data from African, European and Hispanic ancestries. There is no clear evidence that the GRS2 is missing many variants when comparing to AAGRS which is based on African ancestry data. However, the distributions of the scores between ancestries can be very different. The consequences of this are that we need to be careful about combining ancestries as we may draw wrong conclusions about quality of models or about particular risk to individuals. In addition, to equalise measures such as sensitivity or specificity would require substantially different risk thresholds to be used.

## Data Availability

Access to SEARCH data can be requested via https://www.searchfordiabetes.org/dspHome.cfm.
Access to YODA data is restricted to associated research universities.

## Funding and Assistance

This study was supported by a grant from the Randox Ltd.

This study was supported by the National Institute for Health and Care Research Exeter Biomedical Research Centre. The views expressed are those of the author(s) and not necessarily those of the NIHR or the Department of Health and Social Care. M.J.R.’s work on this analysis was supported by NIH NIDDK grant R01 DK124395. The YODA study was funded by NIHR Global Health Award (17/63/131).

## Prior Presentation

Parts of this study were presented in abstract, poster and presentation form at the 9^th^ Meeting of the Study Group on Genetics of Diabetes in Exeter, United Kingdom 17^th^-19^th^ April 2024.

