## Supplementary material for "The impact of ancestry on performance of type 1 diabetes genetic risk scores: high discrimination performance is maintained in African ancestry populations, but population specific thresholds may improve risk prediction"

1) **Dataset information**

Table S1 Numbers of non-T1D and T1D for the five populations. The rows labelled Non-T1D and T1D are the data predominantly used in the paper. Non-T1D 1000G is additional population data taken from 1000G, the 1000G African superpopulation participants are used to compare with the three African cohorts in our study, the 1000G European superpopulation is compared with the SEARCH US-European population and the Americans superpopulation in 1000G compared with the US-Hispanics.

|  | Uganda | Cameroon | US-Africans | US-Europeans | US-Hispanics |
| --- | --- | --- | --- | --- | --- |
| Non-T1D | 4778 | 223 | 235 | 125 | 170 |
| T1D | 76 | 68 | 194 | 1109 | 266 |
| Non-T1D 1000G | 661 | 661 | 661 | 502 | 347 |

2) **QC, imputation procedures and imputation quality**

Uganda and Cameroon

Pre-QC steps: four batches merged into one file (all on same genotype array)

QC steps (all in PLINK v1.9): any duplicates removed; sample missingness at 0.05; SNP missingness at 0.02; minor allele frequency at 0.001; Hardy-Weinberg equilibrium at 1e-6; samples failing sex-checks (different genetic to phenotype sex or with ambiguous genetic sex) removed.

Additional checks: principal component analysis (PCA) run with 1000G data and assessment for outliers. Kinship assessed.

Imputation: imputation on TOPMed.

SEARCH (US-Africans, US-Europeans, US-Hispanics)

SEARCH has data from two SNP arrays (MEGA and BROAD).

Separate QC: SNP missingness at 0.05; sample missingness at 0.03; minor allele frequency at 0.005 (MEGA); minor allele frequency at 0.01 (BROAD); Hardy-Weinberg equilibrium at 1e-6 (only on Europeans); check PCA; relatedness.

Combined QC: two SNP arrays merged. Perform SNP missingness at 0.01; identify identical samples; checked PCA;

Imputation: imputation on TOPMed.

Uganda non-T1D

QC steps (all in PLINK v1.9): any duplicates removed; sample missingness at 0.05; SNP missingness at 0.02; minor allele frequency at 0.001; Hardy-Weinberg equilibrium at 1e-6.

Additional checks: principal component analysis (PCA) run with 1000G data and assessment for outliers. Kinship assessed.

Imputation: imputation on TOPMed.

Cameroon non-T1D

QC steps (all in PLINK v1.9): any duplicates removed; sample missingness at 0.02; SNP missingness at 0.02; minor allele frequency at 0.001; Hardy-Weinberg equilibrium at 1e-6.

Additional checks: principal component analysis (PCA) run with 1000G data and assessment for outliers. Kinship assessed.

Imputation: imputation on TOPMed.

For the GRS2 some SNPs have low imputation quality (Rsq<0.7) for Cameroon, Uganda T1D and non-T1D. All SNPs are well imputed for the SEARCH data. Most poorly imputed SNPs have very low minor allele frequency. In Figure S1 the allele frequencies for any poorly imputed SNPs are shown; blue dots are protective alleles, and the red crosses are risk increasing alleles. The size of the markers represents the weight of the SNP in the GRS. Almost all the poorly imputed SNPs have no significant difference in frequencies between T1D and nonT1D data. The Cameroon SNP shown with a MAF of approximate 0.5 (T1D) and 0.3 (non-T1D) has a low weight of 0.22 and an imputation quality of 0.67. The effects of poor imputation quality overall are small.

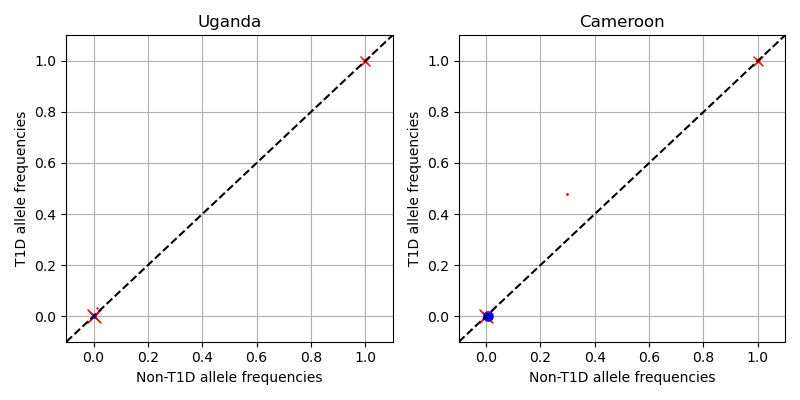

Figure S1 The allele frequencies for any poorly imputed SNPs are shown. The blue dots are protective alleles, the red crosses are risk increasing alleles. The size of the markers represents the weight of the SNP in the GRS

3) **Discriminative power of GRS1, GRS2 and AAGRS on the datasets**

Table S2 AUCs and uncertainties for the five populations and three GRSs. Uncertainties are at 95% level. These are tabular results of the bottom right plot of Figure 1 of the main paper.

|  | **AUC** | | |
| --- | --- | --- | --- |
| **Population** | **GRS1** | **GRS2** | **AAGRS** |
| Uganda & Cameroon | 0.816 (0.772-0.857) | 0.882 (0.845-0.914) | 0.874 (0.838-0.907) |
| Uganda | 0.800 (0.741-0.855) | 0.890 (0.848-0.926) | 0.879 (0.834-0.917) |
| Cameroon | 0.834 (0.776-0.888) | 0.876 (0.819-0.924) | 0.885 (0.835-0.929) |
| US-Africans | 0.796 (0.751-0.837) | 0.839 (0.799-0.877) | 0.838 (0.799-0.874) |
| US-Europeans | 0.840 (0.799-0.876) | 0.878 (0.841-0.912) | 0.810 (0.764-0.851) |
| US-Hispanics | 0.807 (0.767-0.856) | 0.868 (0.832-0.901) | 0.789 (0.742-0.834) |

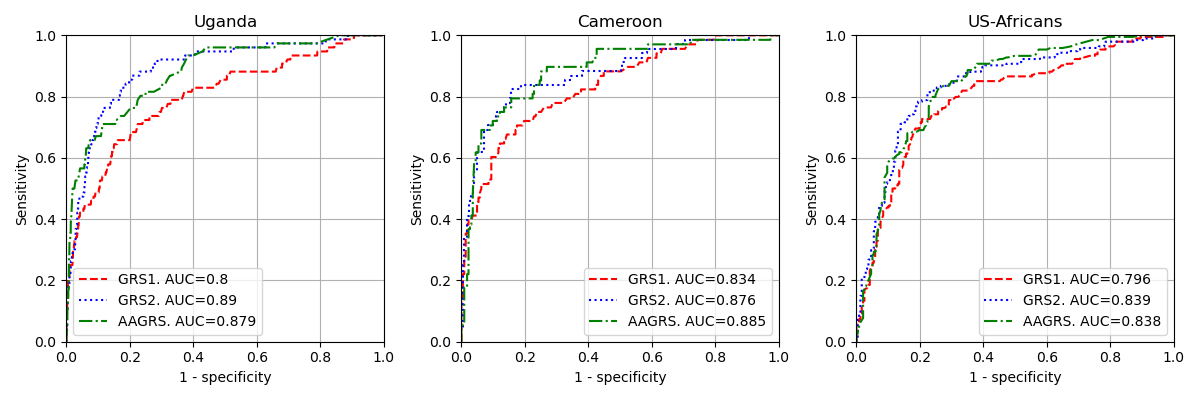

Figure S2 ROC curves for the three GRSs for the separated-out Uganda and Cameroon datasets alongside the US-Africans data. AUCs for the three GRSs are in the legend.

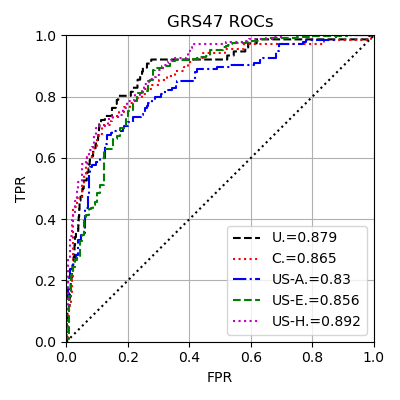

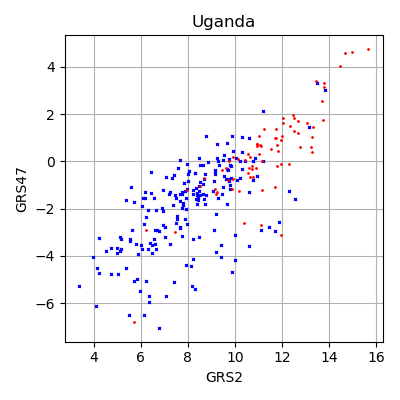

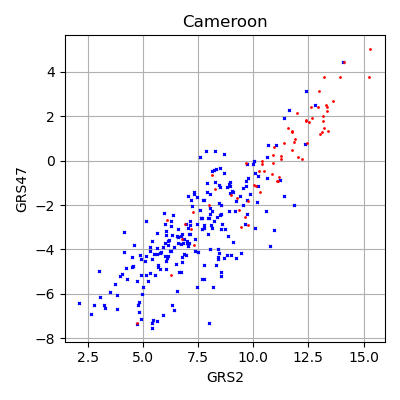

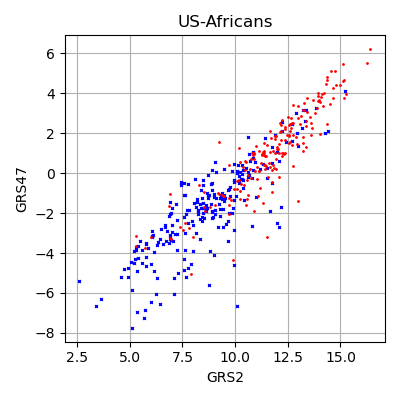

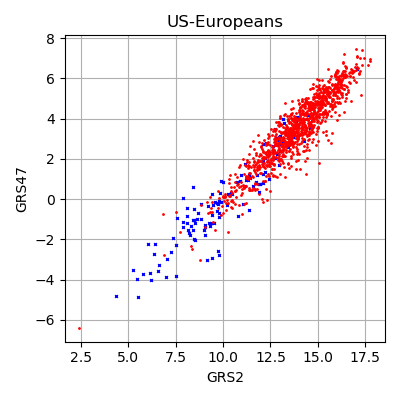

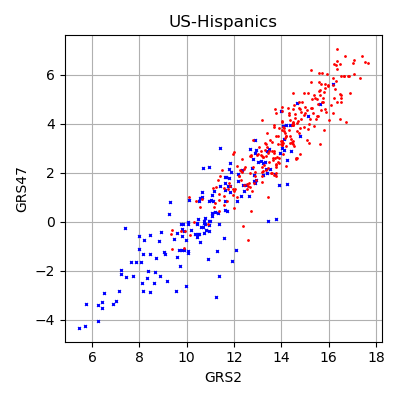

Figure S3 Results for the GRS47. Top left) ROCs and AUCs on the datasets; AUCs in the legend. Rest of plots) Direct plots of the GRS47 against the GRS67. For Uganda the number of non-T1D shown is a random 200.

Table S3 GRS2 AUCs when using 1000G data as the non-T1D data.

|  | **GRS2** |
| --- | --- |
| **Uganda** | 0.871 (0.824-0.912) |
| **Cameroon** | 0.811 (0.742-0.872) |
| **US-Africans** | 0.877 (0.845-0.908) |
| **US-Europeans** | 0.904 (0.887-0.919) |
| **US-Hispanics** | 0.883 (0.856-0.908) |

4) **GRS1, GRS2 and AAGRS distributions on the datasets**

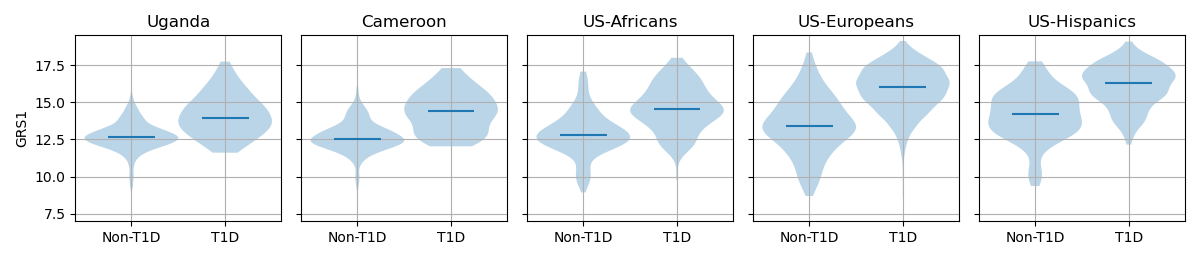

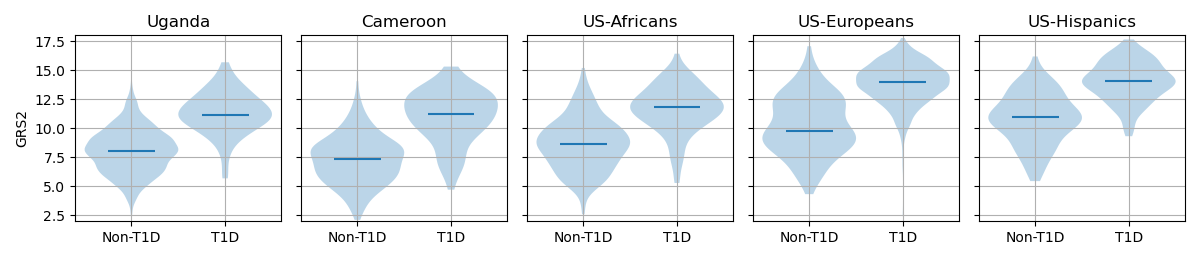

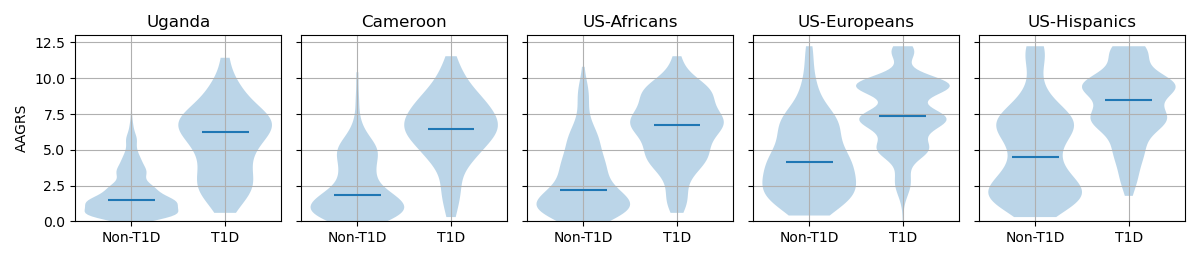

Figure S4 Separated out GRS distributions for the Uganda and Cameroon data for the three GRSs.

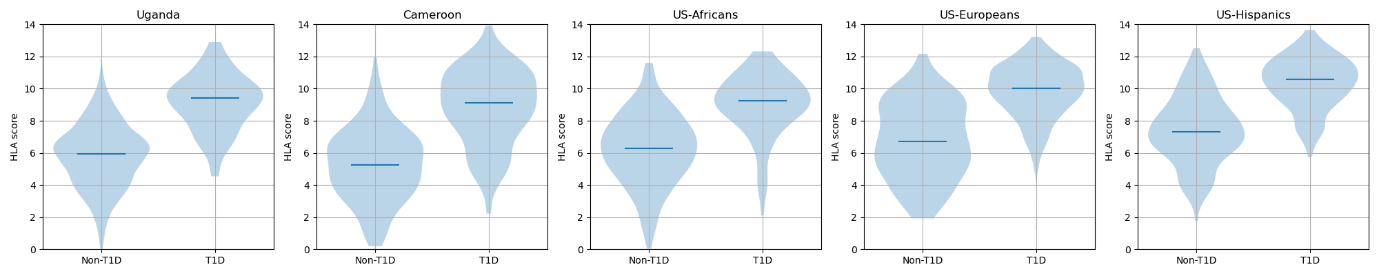

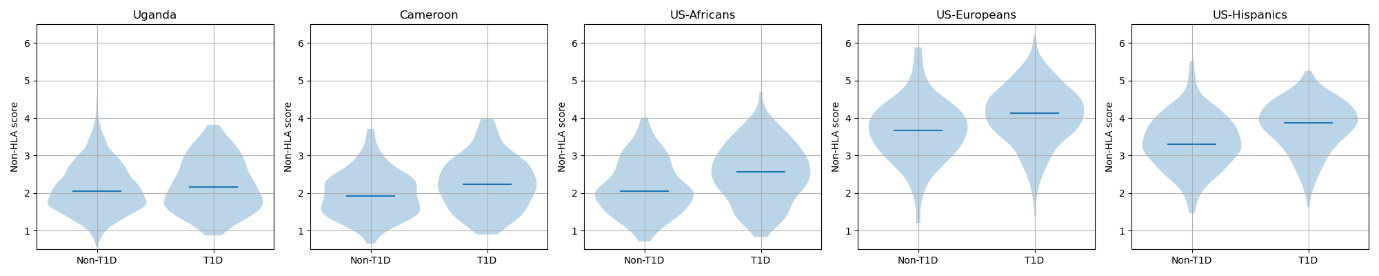

Figure S5 Distributions of the linear scores of the 35 HLA SNPs (top row) and 32 non-HLA SNPs (bottom row) for the five datasets.

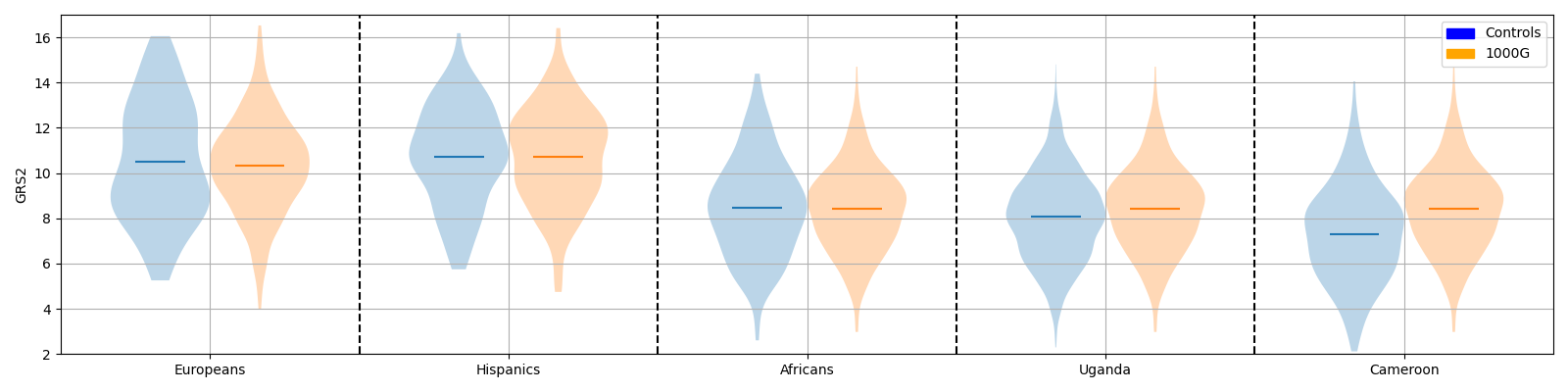

Figure S6 The non-T1D data (labelled as Controls) for each population with the equivalent data from the 1000G dataset.

5) **Sensitivity and specificity with different thresholds**

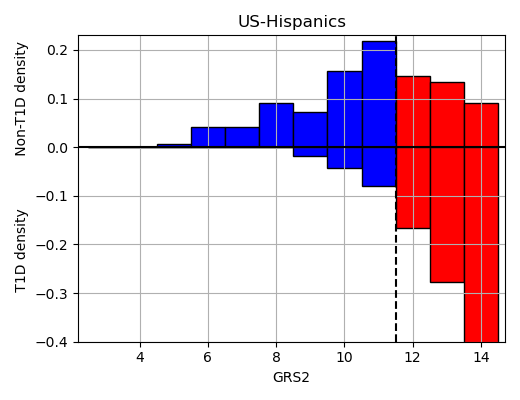

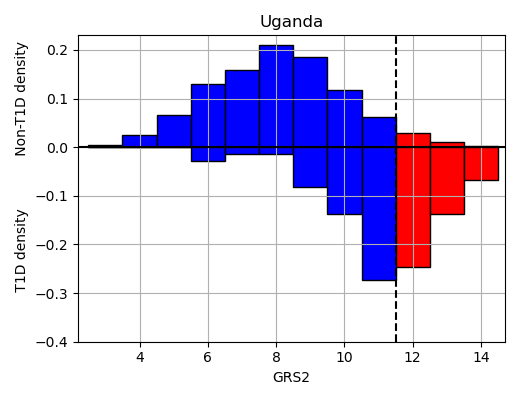

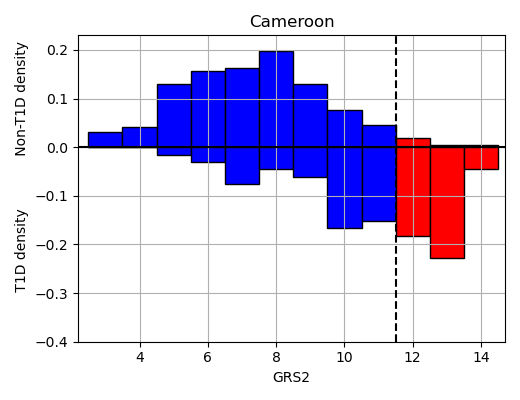

Figure S7: The densities of the non-T1D and T1D data at different GRS values for the US-Hispanics, Uganda and Cameroon populations. Samples to the left of the vertical dashed line would be defined as low risk (in blue columns) and to the right of the line as high risk (red columns). The number of non-T1D and T1D samples are shown as bars above and below the horizontal line respectively.

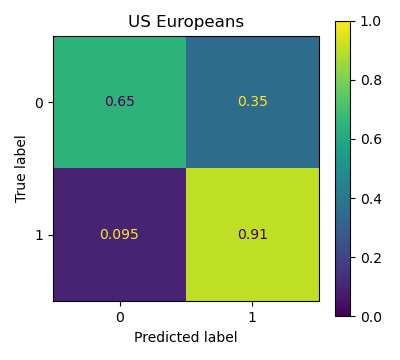

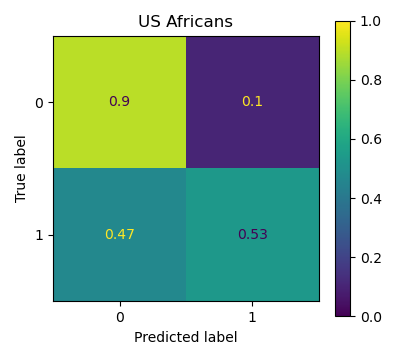

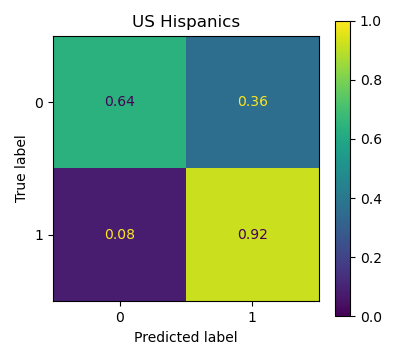

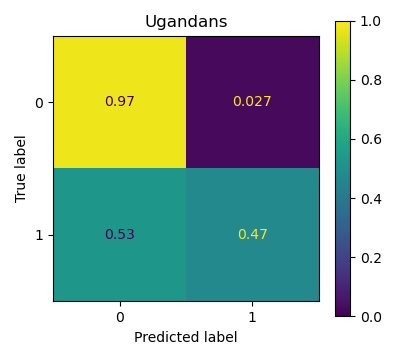

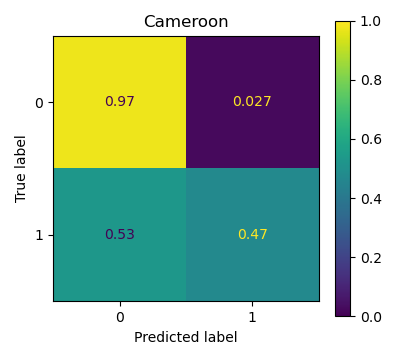

Figure S8. Confusion matrices for US-Europeans, US-Africans, US-Hispanics, Uganda and Cameroon populations with a threshold of 11.5 on the GRS2. These are the confusion matrices related to data shown in Figure 3 of the main paper.

6) **The importance of correctly matching populations for cases and controls**

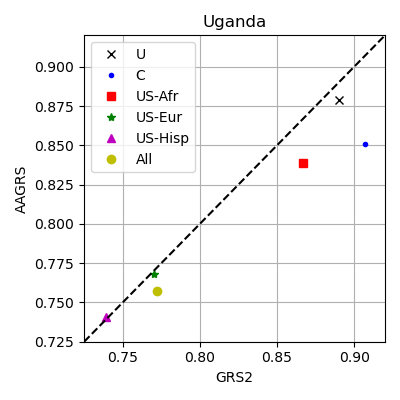

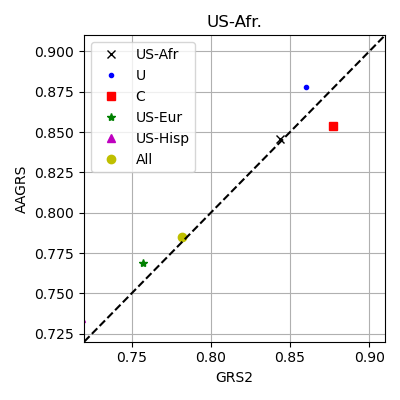

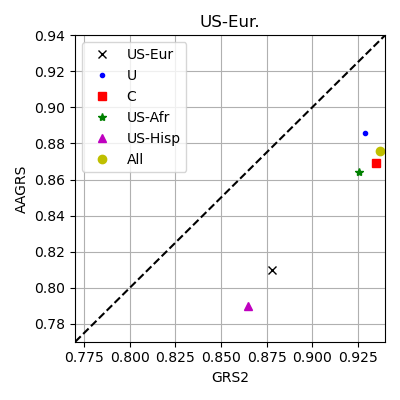

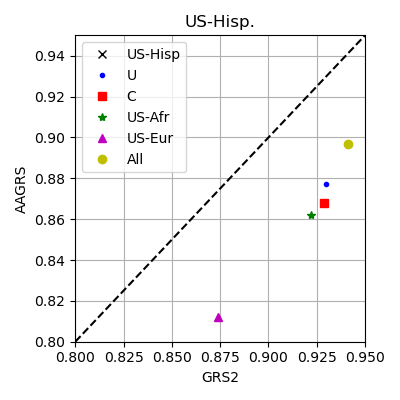

Figure S9 The AUCs for the AAGRS and GRS2 if the non-T1D controls are a combination of populations (with under-sampling so the non-T1D population importance is equalised) for the T1D data from populations other than Cameroon. The black cross is where the non-T1D population is just the specified population, the yellow circle is for all five populations included in the non-T1D populations and the others are pairs of the specified populations with the other populations.

7) **Direct comparisons of GRSs and combining GRSs**

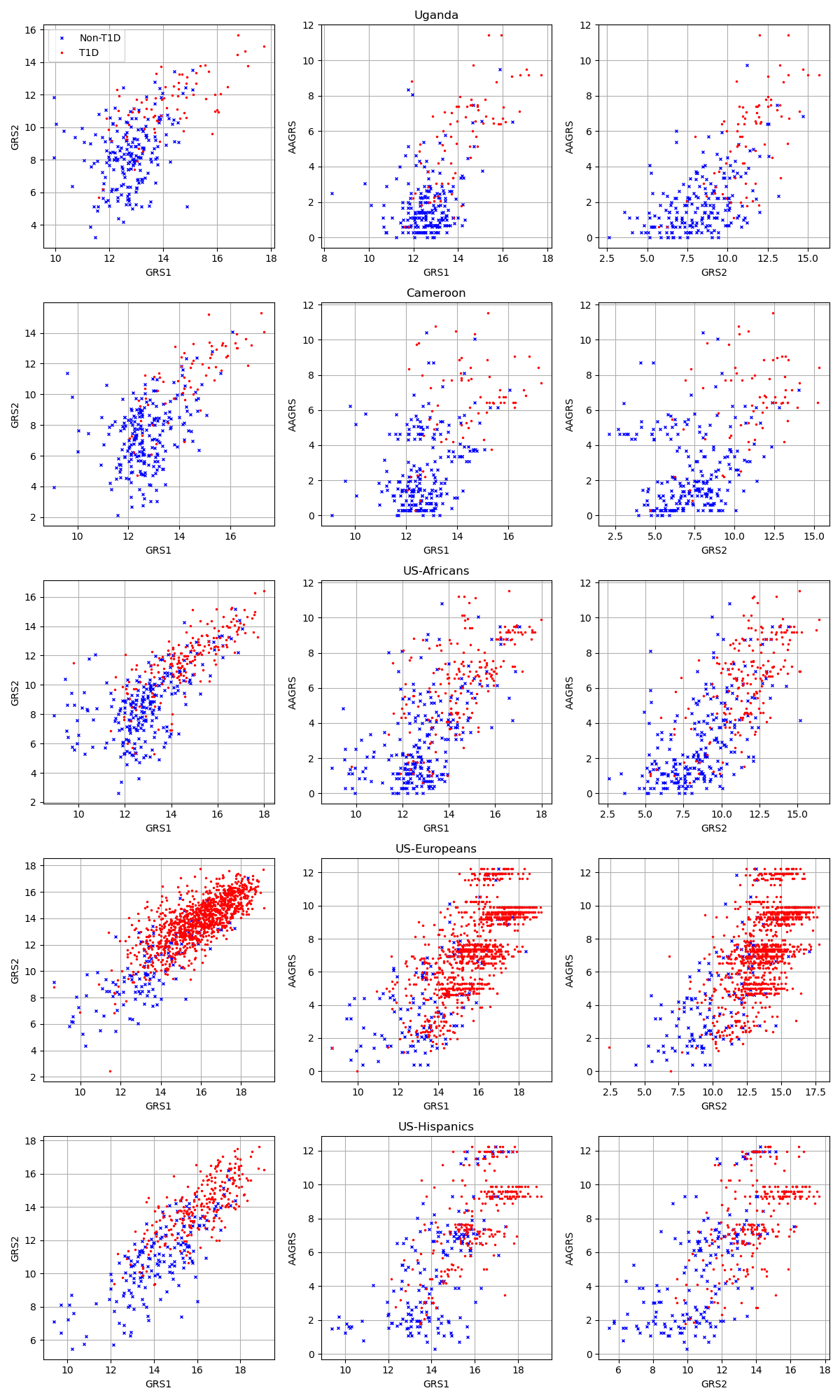

Figure S10 Plots of GRSs against one another. Red markers are T1D and blue markers non-T1D.

Table S4 Spearman rank correlation coefficients between GRSs for the five populations. Confidence intervals shown at the 95% level.

|  | **GRS1-GRS2** | **GRS1-AAGRS** | **GRS2-AAGRS** |
| --- | --- | --- | --- |
| **Uganda** | 0.347 (0.319-0.373) | 0.323 (0.296-0.351) | 0.449 (0.425-0.474) |
| **Cameroon** | 0.574 (0.479-0.657) | 0.510 (0.418-0.593) | 0.485 (0.378-0.584) |
| **US-Africans** | 0.794 (0.748-0.833) | 0.694 (0.638-0.742) | 0.758 (0.714-0.794) |
| **US-Europeans** | 0.780 (0.753-0.804) | 0.739 (0.713-0.762) | 0.646 (0.610-0.680) |
| **US-Hispanics** | 0.832 (0.796-0.860) | 0.754 (0.711-0.791) | 0.723 (0.674-0.764) |

**A**

**C**

**B**

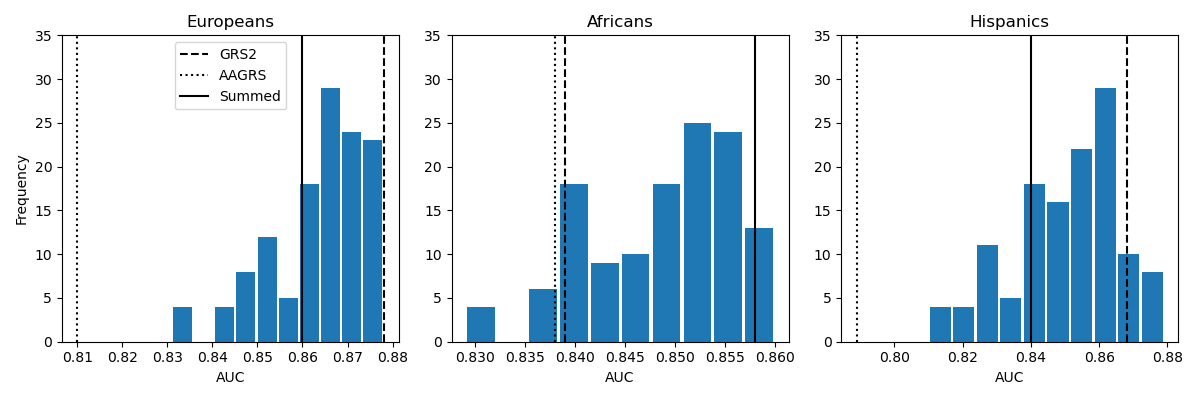

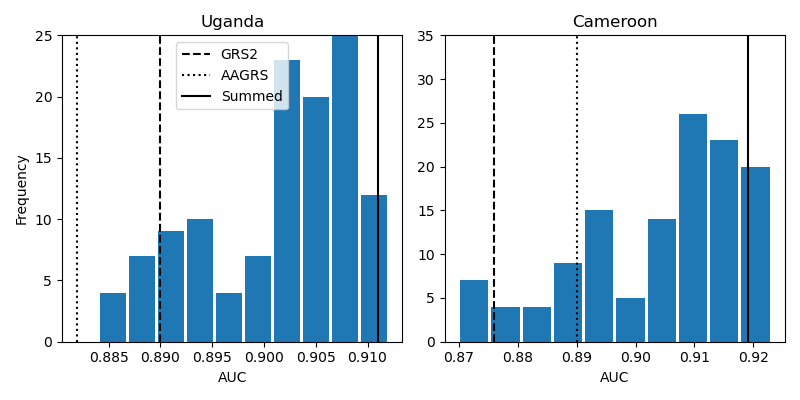

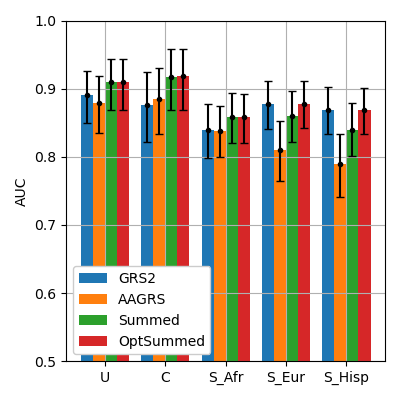

**F**

**E**
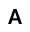

**D**

Figure S11 Summing the GRS2 final score with all possible combinations of the GRS7 SNPs; the weighted scores of all combinations of the GRS7 SNPs are added to the GRS2 and the AUC calculated. These results are displayed as histograms for the five datasets (A-E). And with the optimal weighted sum of the final scores between GRS2 and GRS7 (shown in F); where the weighting that maximises the AUCs between the two final GRSs is used. Top row (A-C) is SEARCH populations.

8) **SNP importance for the AAGRS and GRS2**

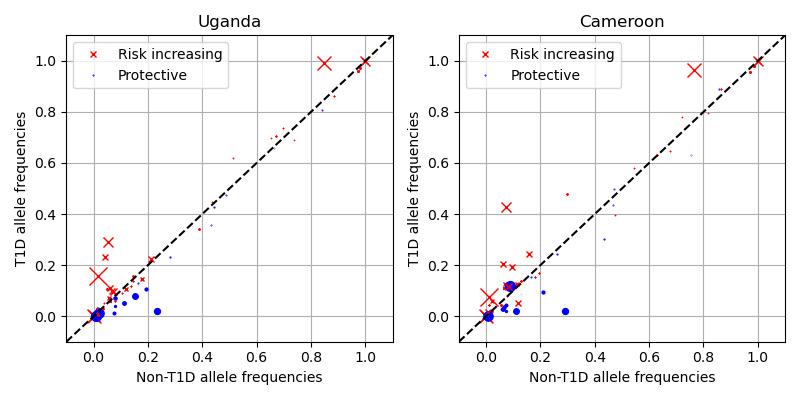

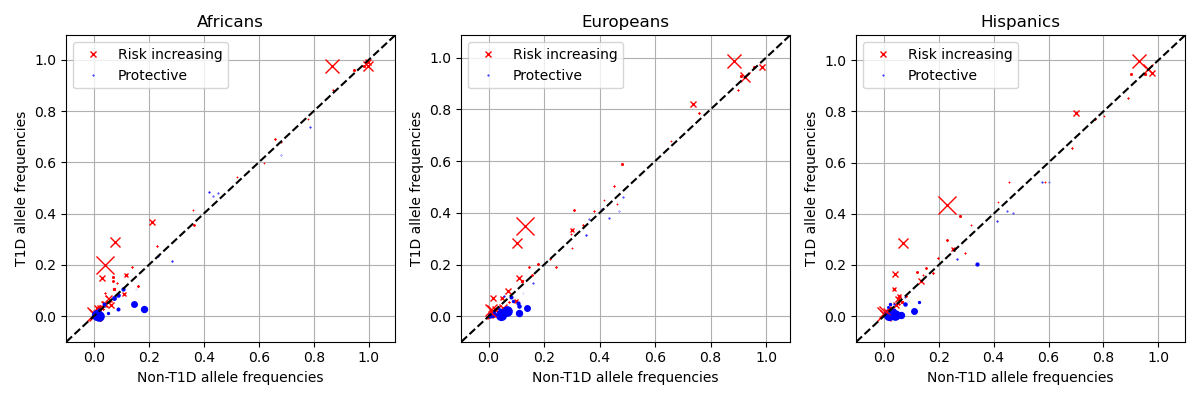

Figure S12 Allele frequencies for the T1D and non-T1D data. The red crosses and blue dots represent risk increasing and protective alleles respectively. The marker size is proportional to the magnitude of the effect score.

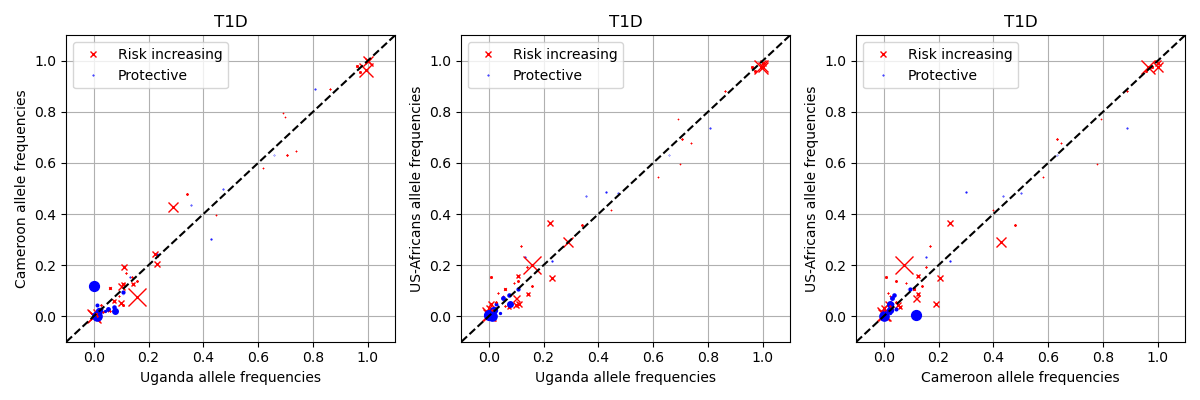

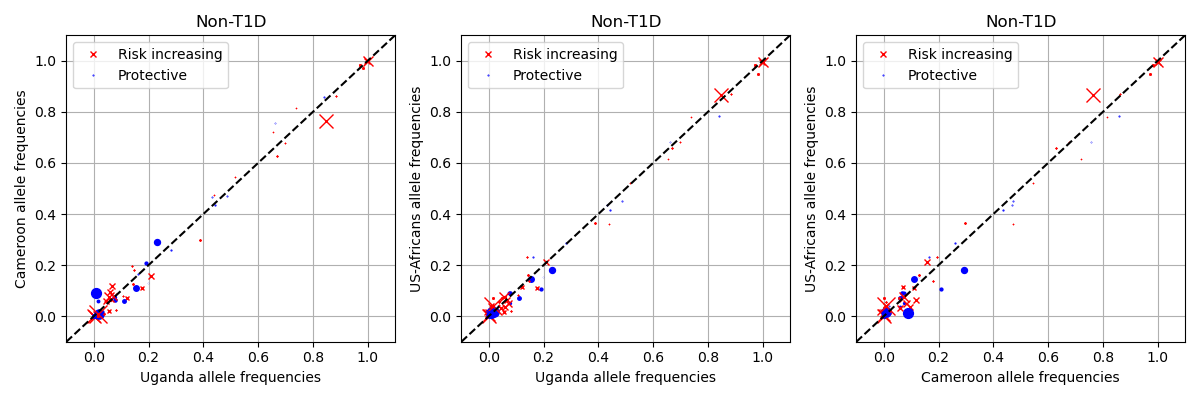

Figure S13 Allele frequencies for the three African ancestry populations compared to one another. The red crosses and blue dots represent risk increasing and protective alleles respectively. The marker size is proportional to the magnitude of the effect score.

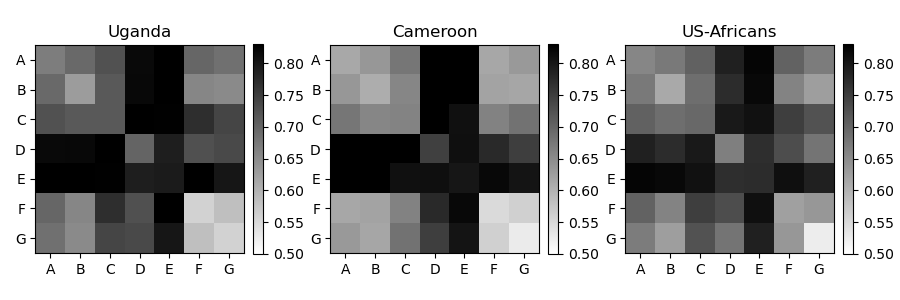

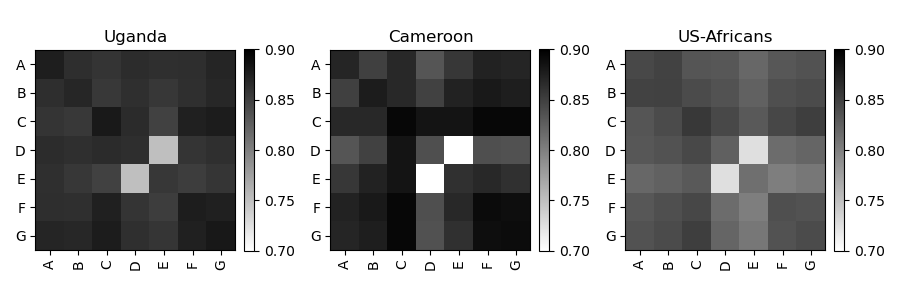

Figure S14 Inclusion (top row) and removal (bottom row) of individual (across the diagonal) and pairs of the AAGRS SNPs. The colours in the plots are the AUC scores when included/excluding those SNPs (with the AUC scale shown at the side). SNPs are denoted as A-G for ease of viewing: A=6:32450613; B=6:32583299; C=6:32591213; D=6:32605884; E=6:32626272; F=11:2182224; G=17:38066240. Chr:pos are from Genome Reference Consortium Human Reference 38. In rs notation: A=rs34303755; B=rs34850435; C=rs9271594; D=rs2187668; E=rs9273363; F=rs689; G=rs2290400.

Table S5 Mean weighted SNP differences between T1D samples and non-T1D samples; any SNP with over 0.1 difference. Chr:pos are from Genome Reference Consortium Human Reference 38.

| Chr:pos (HG38) | RSID | Uganda | Cameroon | Africans | Europeans | Hispanics | Mean |
| --- | --- | --- | --- | --- | --- | --- | --- |
| chr6:32705608:C:G | rs9275490 | 0.596 | 0.268 | 0.66 | 0.914 | 0.855 | 0.659 |
| chr6:32658707:T:C | rs9273369 | 0.594 | 0.89 | 0.538 | 0.463 | 0.548 | 0.607 |
| chr6:32644083:A:G | rs9273032 | 0.606 | 0.769 | 0.447 | 0.272 | 0.175 | 0.454 |
| chr6:32615766:A:G | rs9271347 | 0.494 | 0.671 | 0.361 | 0.343 | 0.226 | 0.419 |
| chr6:32635435:T:C | rs9469200 | 0.195 | 0.232 | 0.258 | 0.274 | 0.236 | 0.239 |
| chr6:32479411:T:A | rs9269173 | 0.251 | 0.195 | 0.158 | 0.076 | 0.168 | 0.17 |
| chr11:2159830:T:G | rs3842753 | 0.023 | 0.139 | 0.255 | 0.141 | 0.161 | 0.144 |
| chr6:33077179:T:C | rs6934289 | 0.118 | 0.154 | -0.001 | 0.069 | 0.181 | 0.104 |
| chr6:32704437:G:T | rs62406889 | 0.023 | 0.03 | 0.077 | 0.199 | 0.161 | 0.098 |
| chr6:29840255:C:G | rs1233320 | 0.075 | 0.14 | 0.017 | 0.012 | -0.003 | 0.048 |
| chr6:32680817:A:G | rs28746898 | 0.034 | -0.132 | 0.034 | 0.186 | 0.079 | 0.04 |

**Correlations between AAGRS and GRS2 SNPs**

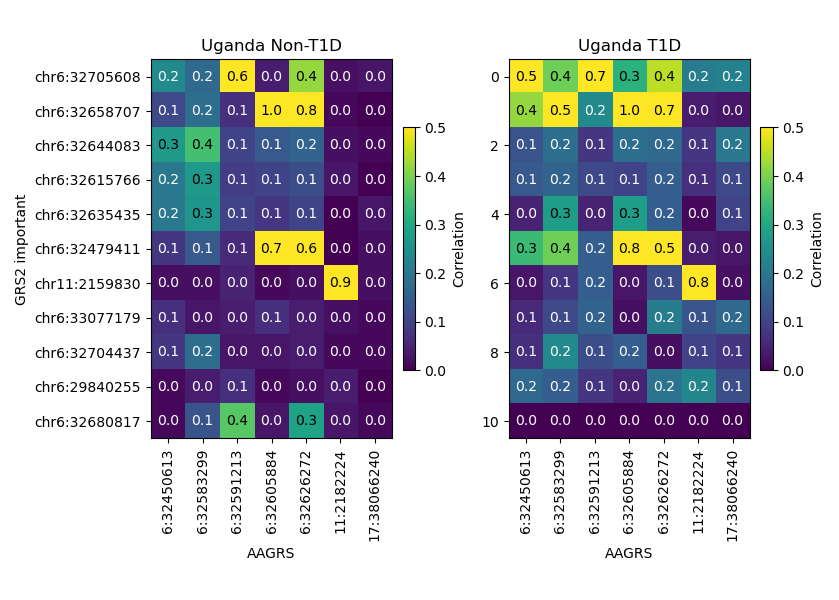

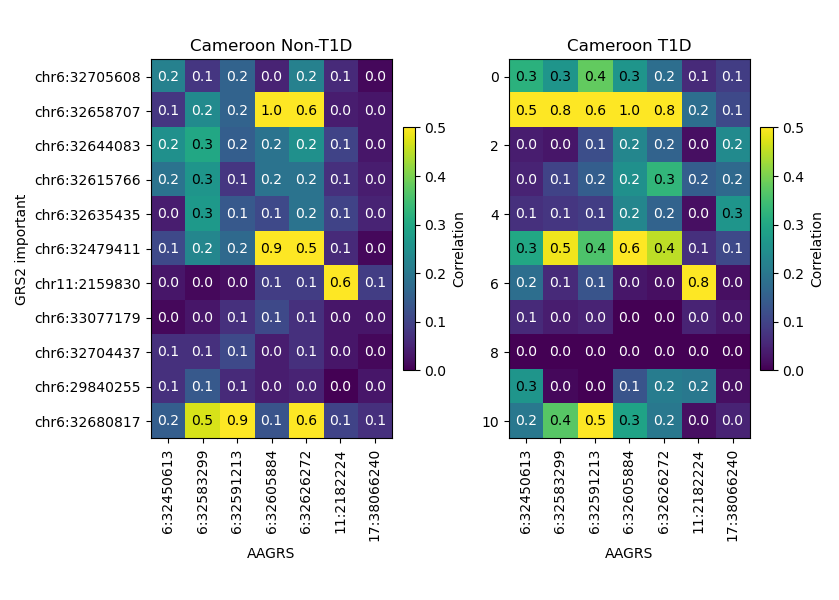

Figure S15 SNP-wise correlations between the GRS7 SNPs and 11 of the GRS2 SNPs. The GRS2 SNPs are selected as any with over a 0.1 difference between T1D and non-T1D; the same as in Table S6. Chr:pos are from Genome Reference Consortium Human Reference 38.
